# Trial-level factors affecting accrual rate of systemic sclerosis randomized clinical trials over 20 years

**DOI:** 10.1101/2024.09.26.24314451

**Authors:** Barbara Russo, Iulia-Simona Chirică, Delphine Sophie Courvoisier, Michele Iudici

## Abstract

**Objectives:** To estimate the average time to complete patient enrollment and identify factors associated with accrual rates in systemic sclerosis (SSc) randomized controlled trials (RCTs).

**Methods:** We searched published SSc-RCTs indexed in PubMed from 2000 to 2024, selecting those with recruitment completed before the COVID-19 pandemic. We recorded key trial features (country, phase, randomization ratio, intervention, blinding, funding source, outcome type) and enrollment year(s). We measured enrollment duration and accrual rate (participants per month). A multivariable negative binomial generalized linear model was used to identify factors associated with accrual rate.

**Results:** We included 80 studies, mostly single-country (75.0%) and industry-funded (57.5%), mainly recruiting in Europe (36.2%) and North America (22.5%). In 65% of studies, both limited and diffuse SSc patients were enrolled. The median sample size was 40.5 patients, with 20% of RCTs enrolling ≥100 patients. The median recruitment time was 15 months (IQR 9.9 – 30.0), with a median accrual rate of 3.1 (IQR 1.6 - 5.5) participants per month. Recruitment rates varied over time, with faster accrual early in the 2000s and after 2012, and a slower period in between. Multivariable analysis showed that accrual rate was positively associated with skewed randomization, non-industry funding, international recruitment, and inclusion of both SSc subsets, especially compared to studies involving only dcSSc patients.

**Conclusions:** Recruiting SSc patients for RCTs has been challenging, with generally slow accrual over the past 20 years and no significant improvement over time.

## Introduction

Randomized controlled trials (RCTs) are essential for understanding the efficacy and safety of medical interventions. However, enrolling participants in RCTs remains a significant challenge (1). A substantial proportion of trials fails to recruit the expected number of patients, or often necessitates longer enrollment periods. This delay increases costs and hinders the timely application of results in clinical practice.

Several factors influence the timely recruitment of patients into clinical trials (2). Slow patient accrual can be unrelated to trial design. Issues such as the rarity of the disease, patient skepticism towards research, a lack of interest or incentive among healthcare providers to recruit participants, language barriers, and the need for patients to travel long distances to the recruitment site can all impair patient recruitment (2). Nevertheless, patient accrual can be negatively affected by specific trial features, such as restrictive eligibility criteria, the selection of surrogate outcome measures, and the lack of blinding (2, 3). It could also be positively affected by using active comparators, employing skewed randomization to increase the likelihood of receiving the treatment arm, and implementing an open-label design, which may all encourage patient participation in clinical trials (4). However, analyses of these factors have yielded inconclusive results (5-8) and this aspect has not yet been studied in the context of rare autoimmune diseases.

Systemic sclerosis (SSc) is a rare connective tissue disease characterized by skin and internal organ fibrosis, leading to high morbidity and mortality (9). SSc is an example of difficult-to-study disease because of the low prevalence, the heterogeneity of clinical picture and disease evolution, and the paucity of efficacious treatment (10). About one-third of SSc-RCTs fails mainly because of poor patient recruitment (1). Together with the analysis of patient-, healthcare provider-, and site/community-related barriers to recruitment, a comprehensive understanding of trial-level factors affecting accrual of SSc-RCTs is pivotal to develop strategies devoted to improve and fasten participation to clinical trials.

Herein, we analyze the average time needed to enroll patients in SSc-RCTs conducted in past years and explore the trial-related factors associated with the pace of patient recruitment.

## Methods

As the study did not concern human or clinical data, we did not record the protocol on PROSPERO. We followed the reporting guidelines for meta-analyses and systematic reviews of randomized controlled trials, Preferred Reporting Items for Systematic Reviews and Meta-Analyses (PRISMA) statement, with the exception of those relevant only to meta-analyses (eg, risk of bias assessment)(11).

### Search strategy

We performed an electronic search of MEDLINE via PubMed on 31 May 2024 to identify RCTs on SSc published since 2000. We used the following combination of free terms and MeSH terms (“CREST”[tiab] OR “Scleroderma, Systemic”[Mesh] OR “Systemic sclerosis”[All Fields] OR “scleroderma”[All Fields]) to identify papers on systemic sclerosis. The Cochrane Highly Sensitive Search Strategy was applied for identifying randomized trials (12).

### Eligibility criteria

#### Inclusion criteria

We included primary reports of SSc-RCTs published since 2000 whose recruitment was completed before the start of COVID pandemic (February 2020) to ensure that the accrual rate was not affected by COVID. We defined RCT as a clinical study randomly allocating participants to different interventions.

#### Exclusion criteria

Studies including patients with scleroderma-like disorders such as morphea, localized scleroderma, or other scleroderma-like diseases (GVHD, toxic-related, etc); secondary publications of RCTs (open-label extension, post-hoc analysis); nonrandomized studies; observational studies; meeting abstracts; studies not in English language or published before 2000. We put no restriction for treatment, outcome, or study phase.

### Data collection

All retrieved references were downloaded in the free online program Rayyan (Qatar Computing Research Institute, https://www.rayyan.ai), a systematic review web-based application. Two researchers (ISC, BR) independently checked each title and abstract to exclude irrelevant papers. The full text article was retrieved to confirm eligibility if information in the abstract was unclear or insufficient. The same reviewers then independently examined full-text articles to determine eligibility. Consensus was reached by discussion in case of disagreement. A third reviewer (MI) was available in case of unsolved disagreement. We documented the primary reason for exclusion of full-text articles.

### Data Extraction and Management

Two authors (ISC, BR) extracted the data using a standardized form, and a third author (MI) checked them for consistency. Consensus was reached by discussion. From each study, the following data were obtained: enrolling country(ies), year of publication, funding (industry-or non-industry funded), study phase, intervention (pharmacologic, non-pharmacologic), number of patients enrolled, year of starting recruitment, SSc subset studied (diffuse cutaneous SSc, limited cutaneous SSc, both), complication investigated (skin, lung, Raynaud’s phenomenon/digital ulcers, gastrointestinal tract, other).

Variables: The primary outcome of our analysis was the patient accrual rate, calculated by dividing the number of participants enrolled in the study by the number of months of recruitment (enrolment period). We chose this measure because it represents the efficiency of participant recruitment and is consistent with prior attempts to study accrual (7). Data on the enrolment period were searched throughout the full text (including tables and figures) and supplementary files. If not reported, we checked online trial registration repositories (e.g., WHO International Clinical Trials Registry Platform, ClinicalTrials.gov). The recruitment period was defined as the interval between the enrolment start date and the date of recruitment of the last patient. If the enrolment date of the last patient was not reported, we estimated the recruitment period as the time span between the trial start date and the study completion date - when the last participant was examined or received treatment to gather final data for primary and secondary outcomes - then deducted the time needed to assess the outcomes from this interval.

Information about the enrolling country(ies) was obtained as follows: first, we checked whether the recruitment centers were reported in the full text; if no explicit information was available, we looked at the location of the institutions linked to the authors; if all the institutions were from the same country, we considered the study to have taken place in that country; in case the authors came from different countries and if information about recruiting centers was lacking in the full-text, we checked the supplementary online material, the published protocols, or the trial registration online repositories (e.g. WHO - International Clinical Trials Registry Platform, ClinicalTrials.gov). If the authors were from institutions in different countries/continents, we considered the study to be international/intercontinental.

A study was considered being industry-funded if the sponsor or one of the collaborators was industry. For each RCT, we collected primary outcome(s). Each primary outcome was then independently classified by 2 of the authors (BR and MI) as ‘patient important’ or ‘surrogate’ outcomes according to previous works on this topic (10, 13)). We classified patient-important outcomes as measures that directly impact on quality of life, such as major morbid events (e.g., end-stage lung disease, loss of hand function, etc) or minor morbid events (e.g., pain and functional status); surrogate outcomes were classified as measures that may indicate disease progression and increased risk for patient-important outcomes, or as assessed response to physiologic or laboratory testing without direct tangible effects on patients (e.g., capillaroscopic pattern, worsening of a respiratory parameter, etc.) (10, 13). In case of disagreement, consensus was reached by discussion.

## Data analysis

Data were summarized as number (percentage) for qualitative variables and median (interquartile range) for continuous variables. Continuous variables were compared with Student’s t-test, Wilcoxon test, or Kruskal-Wallis test, and categorical variables with chi-square test or Fisher’s exact test, as appropriate. The outcome variable patient accrual rate was non-normally distributed. We used a multivariable negative binomial generalized linear model with number of patients recruited as the outcome, and the logarithm of the duration of recruitment as offset to assess the impact of trial-level factors previously shown to impact the accrual rate.

The retrieval of the data and their analysis were performed using R 4.4.1 statistical software (R Development Core Team, Vienna, Austria). A P value ≤ 0.05 was considered significant. Ethical approval was not required (study not involving human participants).

## Results

Among the 1160 RCTs identified (flow-chart shown in Figure 1), we included 80 articles, mostly single-country (n = 60; 75.0%), non-industry funded (n = 31; 42.5%), double-blinded (n = 57; 71.3%) studies, having started recruitment between 2011 - 2020 (n = 49; 62.0%), mostly in Europe (n = 29; 36.2%) or North America (n = 18; 22.5%). Most of international studies were funded by industry (n = 17/20). Studies mainly investigated pharmacologic treatments for Raynaud’s phenomenon / digital ulcers (n = 27; 33.8%) or skin involvement (n = 17; 21.2%) and 65.0% included both SSc subsets (limited and diffuse SSc). The median (IQR) sample size was 40.5 patients (24.0 - 87.3), with 16 RCTs (20.0%) having recruited ≥100 patients. We considered ‘patient-important’ the majority of study primary outcome (n = 56; 70.9%).

**Figure 1.**
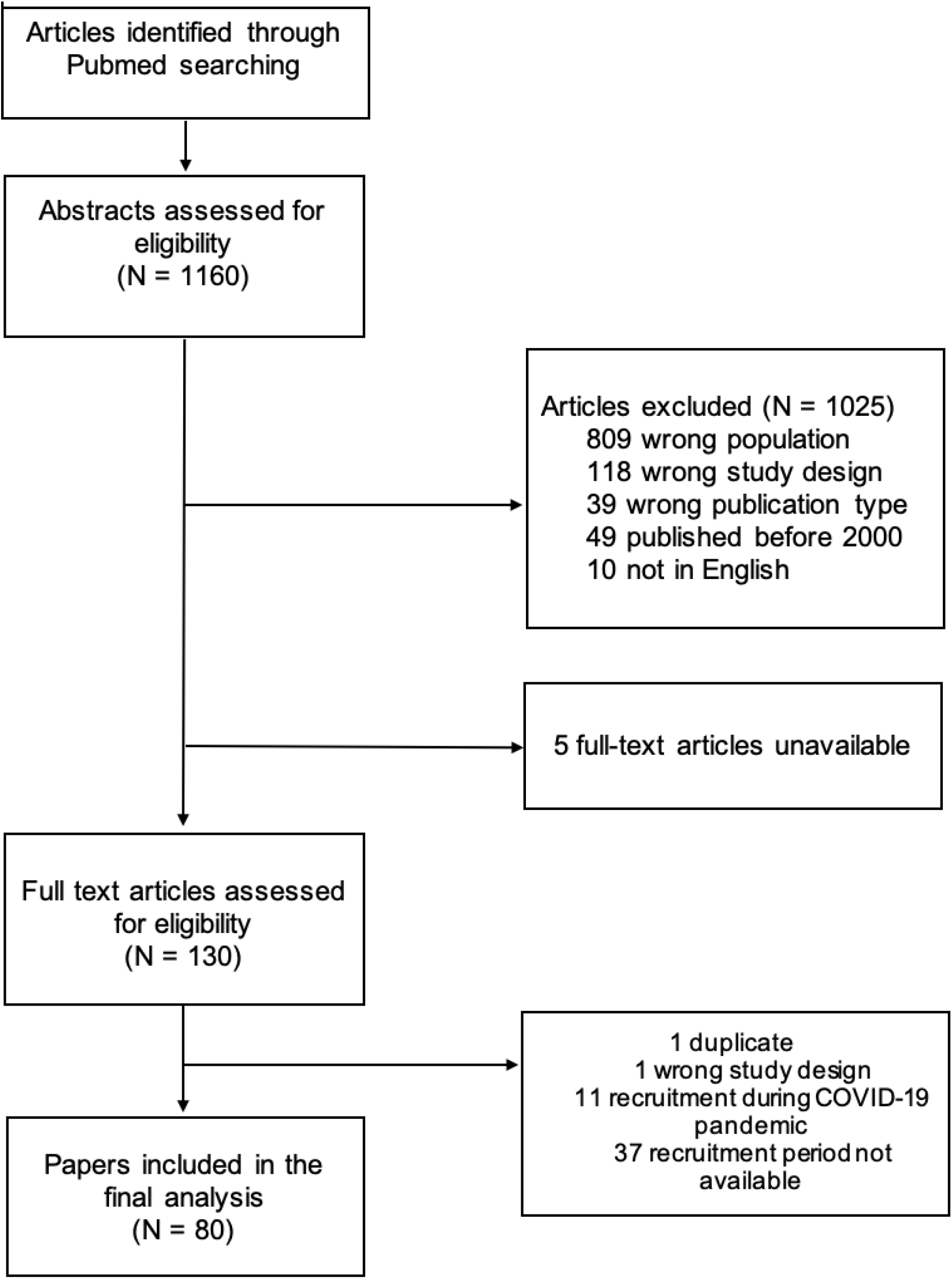
Flow-chart of search strategy.

The median interval duration for participant accrual into the study was 15 months (IQR 9.9 – 30.0), with a median accrual rate of 3.1 (IQR 1.6 - 5.5) patients per month. The unadjusted analysis showed no statistically different accrual rate according to the recruitment period (trials starting before or after 2011), the funding (industry vs. non-industry or jointly funded), the SSc subset enrolled (limited vs. diffuse vs. both), the organ complication studied, the type of intervention (pharmacologic versus non-pharmacologic), the blinding, and the type of primary outcome (patient-important vs. surrogate) (Table 1). A higher accrual rate was found for phase 3 studies recruiting in more continents or being international, and having a skewed randomization.

**Table 1.**
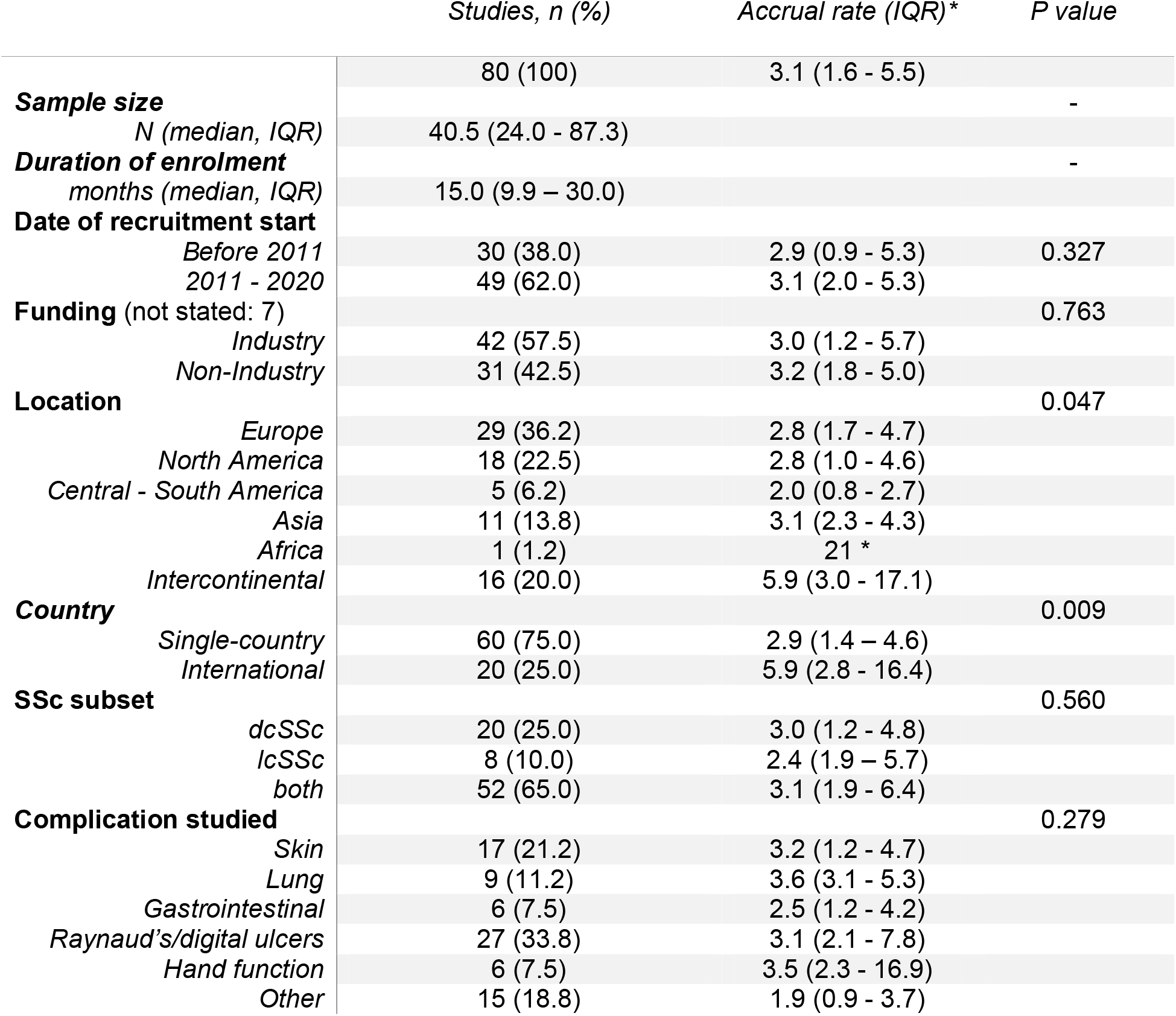
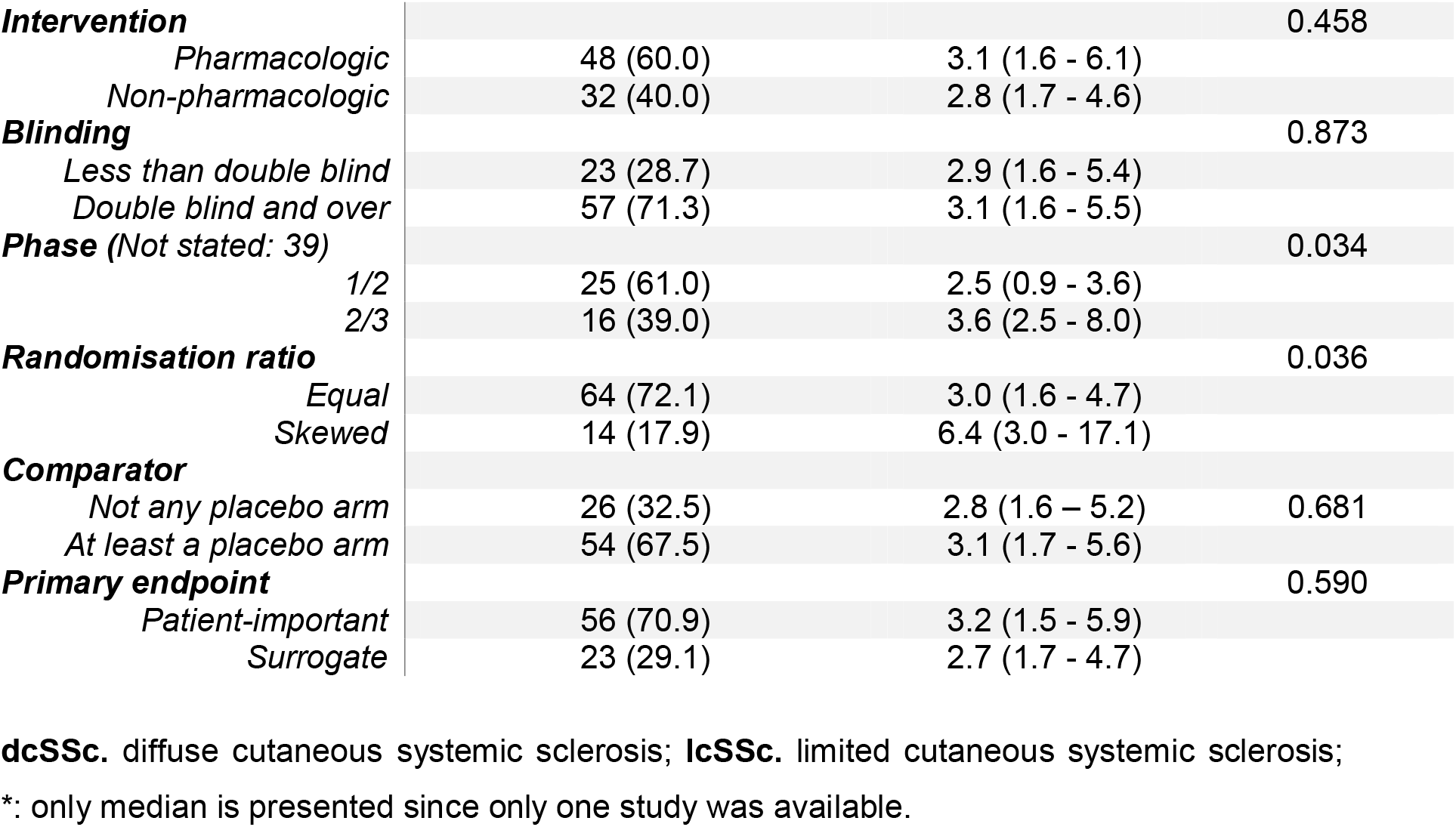
Descriptive characteristics and accrual rate among systemic sclerosis clinical trials included.

Accrual rate changed over time with faster recruitment at the beginning of the 2000 decade, and after 2012, with a period of relatively slow recruitment in between (Figure 2). This trend seems mostly driven by large international studies (top left panel) including both SSc subtypes (bottom left panel). While some studies funded by industry had high accrual rate at the beginning of 2000, recent studies with faster recruitment were equally industry or non-industry funded.

**Figure 2.**
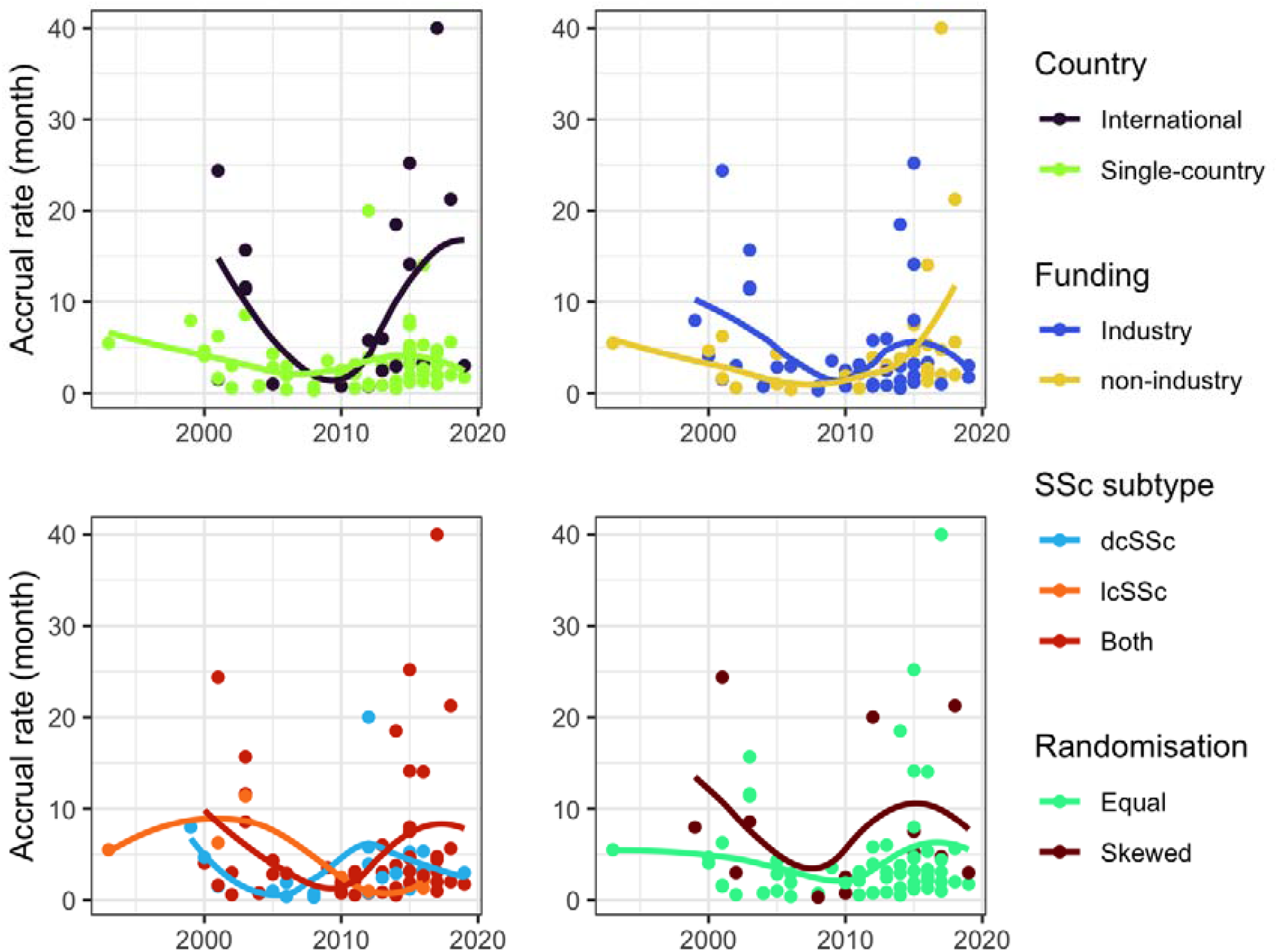
Accrual rate over time by country (top left), funding (top right), SSc subtype (bottom left), and randomisation (bottom right)

In the multivariable analysis, accrual rate was positively associated with a skewed randomisation method, not being funded by industry, and international recruitment. It was also higher in studies including both SSc subsets, especially compared to studies including only dcSSc (Table 2).

**Table 2.** Incidence rate ratio of accrual rate among systemic sclerosis clinical trials included.

|  | *IRR* | *95% confidence interval* | *P value* |
| --- | --- | --- | --- |
| **Blinding (ref: double blind)** | 0.781 | (0.356 – 1.693) | 0.499 |
| **Primary endpoint (ref: patient important)** | 1.229 | (0.790 – 1.946) | 0.337 |
| **Randomisation method (ref: equal)** | 1.870 | (1.114 – 3.295) | 0.016 |
| **Country (ref: single country)** | 3.727 | (0.173 – 0.408) | <0.001 |
| **Funding (ref: industry)** | 1.917 | (1.208 – 3.049) | 0.004 |
| **Comparator (ref: no placebo)** | 1.177 | (0.572 – 2.377) | 0.642 |
| **SSc subset** |  |  |  |
| *both* | -- |  |  |
| *dcSSc* | 0.498 | (0.307 – 0.819) | 0.003 |
| *lcSSc* | 0.766 | (0.421 – 1.513) | 0.406 |

## Discussion

Herein, we sought to examine factors associated with accrual rate in a sample of SSc-RCTs conducted since 2000. On average, trials enrolled 3 patients per month, reflecting an overall slow recruitment. We found that non-industry funding, skewed randomisation, international/intercontinental enrolment, and the inclusion of both SSc subsets were the main features associated with faster enrolment. Interestingly, some of these trial-level factors were more or less present over time, potentially explaining the low accrual rate between 2003 and 2012. Conversely, many factors potentially encouraging trial participation failed to demonstrate association in multivariable analysis. Comparator, type of endpoint, and blinding were not associated with the accrual rate.

In our study sample, the mean duration of the enrollment period was 15 months. The recruitment pace was double (6 patients per month) for trials conducted in multiple countries or continents. However, this rate remains relatively low compared to other conditions. For instance, clinical trials leading to FDA approval of new cancer drugs typically enroll an average of 18 patients per month (8). Even ultra-rare orphan cancer trials have higher recruitment rates than those observed in SSc, with an average of 8 patients per month trials (8). This finding, which further highlights the challenges of conducting RCTs in rare diseases like SSc (14), underscores the need to explore alternative approaches for designing and implementing RCTs in such contexts (15).

Consistent with previous data, we found that patient enrolment per month was higher for larger studies recruiting across different countries or continents (3, 5, 6, 16). We did not find any difference between trials conducted in Europe, North America, or Asia. Notably, industry-funded studies on SSc did not exhibit a higher accrual rate. In fact, the multivariable model indicated a lower accrual rate after adjusting for international settings. Exploring whether this is due to factors such as SSc patients’ lower willingness to participate in industry-sponsored trials as already shown in other papers (17), the increased complexity of industry protocols(18), suboptimal recruitment site selection (19), or a lack of scientific incentives for recruiters would be an interesting topic for future research in the field. We did not analyze the impact on the pace of recruitment according to the income of recruiting countries, but the studies analyzed were mostly conducted in high-income countries (North America and Europe). Increasing trial participation in low-income countries could accelerate recruitment and enhance the generalizability of study results (20).

Our findings align with previous research that found no association between certain trial features, which are theoretically expected to speed up recruitment, and the accrual time (5-8). Similarly, in line with our results, a study analyzing a large sample of oncology trials reported that neither trial blinding nor the choice of comparator significantly impacted patient accrual (5). Conversely, we found that skewed randomization ratios (2:1; 3:1, etc), which increases patients’ chances of being assigned to the tested intervention, favored faster recruitment. We speculate that the lack of observed influence of the comparator on trial accrual may be due to the fact that the majority of SSc-RCTs (about two-thirds) included at least one placebo arm, given the limited availability of effective treatments for SSc. Nevertheless, offering patients a greater chance of receiving an active treatment might enhance their willingness to participate in the trial. As a caution, since skewed randomization requires a larger patient population, factors such as disease prevalence, feasibility, and costs should be carefully evaluated during the trial design phase (6, 8). Finally, the use of patient-important endpoints instead of surrogate endpoints did not lead to faster accrual among SSc patients, likely because endpoints are rarely discussed with or fully understood by patients (21).

We did not observe any improvement in recruitment performance over time. While our data show a slight decrease in accrual time between 2003 and 2012, overall, there has been no significant change in the pace of accrual from 2000 to the present. The findings of Brøgger-Mikkelsen et al. (19) were consistent with the results of our study. In their analysis of more than 5,000 phase 3 clinical trials registered at ClinicalTrials.gov between 2008 and 2019 (19), the authors revealed a significant increase in recruitment duration, rising from an average of 13 months in 2008 – 2011 to 18 months in 2016 – 2019. Concurrently, the number of participants enrolled in clinical research has decreased significantly between 2012 – 2015 and 2016 – 2019, highlighting that recruitment strategies is less effective today compared to several years ago. This could be explained by the increasing complexity of study procedures (18, 22), which is negatively correlated with patients’ willingness to enter and complete clinical trials (17). To address this issue, we have established an international board comprising researchers, stakeholders, and patients to more effectively identify the main barriers to clinical trial participation for individuals with SSc at the patient, physician, and recruitment site levels.

Our study has some limitations. Our sample is limited to studies that have successfully accrued patients and whose results have been published. We did not analyze factors associated with accrual rate for unpublished trials or for those failing to attain the desired sample size. Future research might focus on a comparison of completed versus terminated trials. Furthermore, we did not analyze whether competing trials could have affected accrual of patients. However, given the rarity of the disease and the geographical dispersion of trials conducted over the years, we think that this issue is negligible in this context. Finally, the inclusion of RCTs published only in journals indexed in PubMed could have led to exclude studies indexed in other search engines. Readers should be aware of this limitation when interpreting the results of the present study.

This work has several strengths. For the first time, we have analyzed how trial-level factors influence the pace of patient enrollment in a rare systemic autoimmune disease such as systemic sclerosis. We hope this analysis will help clinical researchers better estimate the required enrollment time, thereby reducing the risk of failing to achieve the desired sample size or needing to prolong the recruitment period.

In conclusion, enrolling SSc patients for clinical trials is challenging, and on average, recruitment has been relatively slow over the past two decades, with no improvement over time. This study further reinforces the idea that harmonizing efforts across countries to enhance trial recruitment and advance medical knowledge in this field is a priority.

## Data Availability

All data produced in the present study are available upon reasonable request to the authors

## Acknowledgements

None.

## Ethics approval

Not needed.

## Contributors

BR, ISC, MI contributed to planning and data collection, data quality control, data analysis, and interpretation. They drafted and revised the manuscript critically for important intellectual content and gave final approval of the version to be published. BR, ISC, DSC, MI contributed to data collection, data quality control, reviewed the manuscript, and provided important intellectual content. DSC and MI had full access to the study data. All authors have read, revised, and approved this manuscript and had final responsibility for the decision to submit for publication.

